# ‘Inflammaging’ in Major Depressive Disorder: Associations between markers of inflammation and brain ageing in depression across two large community-based cohorts

**DOI:** 10.1101/2021.12.17.21267979

**Authors:** Claire Green, Marco Squillace, Anna J. Stevenson, Aleks Stolicyn, Mathew A. Harris, Ellen V. Backhouse, Emma L. Hawkins, Adele M. Taylor, Zoe Morris, Joanna M. Wardlaw, J Douglas Steele, Gordon D. Waiter, Anca-Larisa Sandu, David J. Porteous, James H. Cole, Stephen M. Lawrie, Jonathan Cavanagh, Simon R. Cox, Andrew M. McIntosh, Heather C. Whalley

## Abstract

**Background:** Major Depressive Disorder (MDD) is associated with accelerated ageing trajectories including functional markers of ageing, cellular ageing and markers of poor brain health. The biological mechanisms underlying these associations remain poorly understood. Chronic inflammation is also associated with advanced ageing; however, the degree to which long-term inflammation plays a role in ageing in MDD remains unclear, partly due to difficulties differentiating long-term inflammation from acute cross-sectional measures.

**Methods:** Here, we use a longer-term measure of inflammation: a DNA methylation-based marker of C-reactive protein (DNAm CRP), in a large cohort of individuals deeply phenotyped for MDD (Generation Scotland, GS; *N*=804). We investigate associations between DNAm CRP and serum CRP using linear modelling with two brain ageing neuroimaging-derived phenotypes: (i) a machine learning based measure of brain-predicted age difference (brain-PAD) and (ii) white matter hyperintensities (WMH). We then examine inflammation by depression interaction effects for these brain ageing phenotypes. We sought to replicate findings in an independent sample of older community-dwelling adults (Lothian Birth Cohort 1936, LBC1936; *N*=615).

**Results:** DNAm CRP was significantly associated with increased brain-PAD (β=0.111, p=0.015), which was replicated in the independent sample with a similar significant effect size (β=0.114, p=0.012). There were no associations between the inflammation markers and WMH phenotypes in the GS-imaging sample, however in the LBC1936 sample, DNAm CRP was significantly associated with both Wahlund infratentorial (β=0.15, P_FDR_= 0.006) and Fazekas deep white matter hyperintensity scores (β= 0.116, P_FDR_=0.033). There were no interaction effects between inflammation and MDD in either cohort.

**Conclusions:** This study found robust associations between a longer-term marker of inflammation and brain ageing as measured by brain-PAD, consistent across two large independent samples. However, we found no evidence for interaction effects between inflammation and MDD on any brain ageing phenotype in these community-based cohorts. These findings provide evidence that chronic inflammation is associated with increased brain ageing, which is not specific to MDD. Future work should investigate these relationships in clinical samples including with other inflammatory biomarkers and should furthermore aim to determine causal directionality.

## 1. Introduction

Major depressive disorder (MDD) is the most common mental health condition in the general population and is the primary cause of disability worldwide (Bromet *et al*., 2011; Marcus *et al*., 2012; Friedrich, 2017). MDD has been associated with poor health outcomes including increased risk of mortality, reduced life expectancy (Chesney, Goodwin and Fazel, 2014; Cuijpers *et al*., 2014), increased risk of cognitive decline and Alzheimer’s disease (Speck *et al*., 1995; Chodosh *et al*., 2007; John *et al*., 2019), advanced physical ageing (such as handgrip strength and walking speed) (Lever-van Milligen *et al*., 2017), and increased risk of poor cardiovascular and somatic health (Penninx *et al*., 2013; Hare *et al*., 2014). It has been proposed that the link between depression and poor physical and cognitive health may be explained by accelerated ageing trajectories in those with MDD (Wolkowitz *et al*., 2010). Biological evidence supporting this includes accelerated epigenetic ageing in MDD (Whalley *et al*., 2017; Han *et al*., 2018), increased immunosenescence (Diniz *et al*., 2019) and telomere shortening (Darrow *et al*., 2016; Lin, Huang and Hung, 2016).

Another recently developed promising biomarker of biological ageing is ‘brain-predicted age’ (Cole *et al*., 2018). Here an estimate of an individual’s brain age is derived by applying machine learning algorithms to structural brain MRI data. This estimate can then be compared to an individual’s chronological age. Having an older ‘brain-predicted age’ relative to chronological age has been associated with other markers of ageing such as poorer hand grip strength, decreased walking speed, poorer cognitive performance and also increased risk of death (Cole *et al*., 2018). MDD has also been associated with an older ‘brain-predicted age’ (Han *et al*., 2020), and with a greater number and intensity of white matter hyperintensities (WMH), which are also considered markers of brain ageing processes (Firbank *et al*., 2005; Herrmann, Le Masurier and Ebmeier, 2008; Wang *et al*., 2014; Debette *et al*., 2019; Wardlaw, Smith and Dichgans, 2019).

Despite substantial evidence of accelerated ageing in MDD, biological mechanisms remain unclear. One proposed mechanism is that long-term systemic inflammation accelerates ageing processes (‘inflammaging’) (Franceschi and Campisi, 2014). Long-term inflammation is also associated with age-related diseases such as dementia and cardiovascular disease, and with markers of poor brain health such as brain atrophy and reduced white matter integrity (Satizabal *et al*., 2012; Marsland *et al*., 2015; Gu *et al*., 2017; Walker *et al*., 2017; Furman *et al*., 2019). These age-related diseases are also more prevalent in individuals with MDD who are at increased risk of cognitive decline, dementia, cardiovascular disease, overall mortality and decreased white matter integrity (Byers and Yaffe, 2011; Hare *et al*., 2014; Cherbuin, Kim and Anstey, 2015; Shen *et al*., 2017, 2019).

Several studies have shown that inflammation is implicated in the pathogenesis of MDD and is associated with MDD symptoms, in particular, somatic depressive symptoms (Dantzer *et al*., 2008; Raison *et al*., 2013; Haapakoski *et al*., 2015; Strawbridge *et al*., 2015; Bhattacharya *et al*., 2016; Green *et al*., 2021). Although inflammation is associated with advanced ageing and MDD, the degree to which chronic inflammation plays a role in the relationship between MDD and accelerated brain ageing remains unclear. One major limitation of previous research is that inflammatory processes in MDD and ageing are typically measured using cross-sectional serum biomarkers such as the highly phasic C-reactive protein (CRP), which cannot capture longer-term chronic processes.

Recently, an epigenetic approach has utilised DNA methylation (DNAm) markers of inflammatory activity associated with CRP (Stevenson *et al*., 2020) to create a measure of longer-term inflammation (DNAm CRP). The DNAm CRP score showed greater longitudinal stability compared to the phasic serum measure of CRP and may represent a more stable signature or archive of longer-term inflammation. This DNAm CRP score has also been shown to be strongly associated with markers of poor brain health including widespread reductions in white matter integrity (as indexed by increased mean diffusivity and decreased fractional anisotropy) and demonstrated much greater effect sizes compared to serum CRP in two independent cohorts (Conole *et al*., 2021; Green, Shen, *et al*., 2021).

To our knowledge no previous study has investigated an epigenetic measure of inflammation in relation to brain age or WMH in MDD. Therefore, in the current study we utilised a large cross-sectional imaging cohort (Generation Scotland) in mid-late life with detailed genetic, epigenetic, behavioural and neuroimaging phenotyping to investigate DNAm CRP associations with brain ageing phenotypes in the context of MDD. This study utilised serum and methylation scores of inflammation to investigate their relationships with two markers of brain ageing (i) brain age scores derived from T1-weighted data and (ii) WMH phenotypes. Finally, depression × inflammation interaction effects were investigated to determine the differential relationship of these inflammatory markers and brain ageing in depression. We also sought to replicate findings in an independent sample of older adults from the Lothian Birth Cohort 1936 where equivalent variables were available.

## 2. Methods

### 2.1 Participants

#### Generation Scotland (GS)

Participants included in this study were recruited as part of the Generation Scotland cohort and included a follow-up sample of approximately *N*=1,000 individuals with additional detailed neuroimaging and mental health data (GS-imaging sample). The recruitment process for the GS-imaging sample has been reported in full previously (Navrady *et al*., 2018; Habota *et al*., 2019). The current sample included an unrelated subset of participants (*N*=804) and full details of how this sample was derived have been reported in full previously (Green, Shen, *et al*., 2021; Green, Stolicyn, *et al*., 2021). Demographic data for this cohort are provided in Table 1.

**Table 1:** Participant Demographics.

| **Variable** | **Unit** | **Generation Scotland** | **Lothian Birth Cohort** |
| --- | --- | --- | --- |
| **Participants** | N | 804 | 615 |
| **Age** | Years (Mean, SD) | 59.5 (9.7) | 72.6 (0.7) |
| **Age Range** | Years | 26-84 | 70-74 |
| **Sex** | Males (N) | 336 | 328 |
|  | Females (N) | 468 | 287 |
| **Study Site** | Aberdeen | 412 | - |
|  | Dundee | 392 | - |
| **Body Mass Index** | Mean (SD) | 28 (5.4) | 27.9 (4.4) |
| **MDD** | N | 242 | 26 |
| **Controls** | N | 562 | 589 |
| **Depression Symptoms\*** | Mean (SD) | 4.6 (3.6) | 2.6 (2.2) |
| **Serum CRP Status** | <4mg/L (N) | 654 | 490 |
|  | >= 4mg/L (N) | 150 | 110 |
|  | NA (N) | - | 15 |
| **WMH Volume Total** | mm<sup>3</sup> (Mean, SD) | 330.2 (252.2) | - |
| **WMH Total Count** | Mean (SD) | 8.2 (7.6) | - |
| **Brain Age** | Years (Mean, SD) | 57.2 (11.6) | 73.6 (6.7) |

\*MDD is ascertained by SCID status in the GS-imaging sample and HADS-D in the LBC1936 sample. Depression symptom scores are calculated by total QIDS scores in the GS-imaging cohort and by HADS-D scores in LBC1936.

Ethical approval was formally obtained from the NHS Tayside committee on research (reference 14/SS/0039), and all participants provided their written informed consent.

#### Lothian Birth Cohort 1936 (LBC1936)

For replication analyses, the LBC1936 sample was used (*N*=615) where equivalent variables to those derived in the GS-imaging subsample were available. This cohort is comprised of individuals born in 1936 who were contacted at approximately 70 years of age for detailed cognitive and medical assessment, genetic and epigenetic data collection, and neuroimaging. Full details of the LBC1936 recruitment, clinical assessment, DNAm CRP score calculation and neuroimaging acquisition have been reported in full previously, and the current study used the wave 2 sample with the maximum available imaging sample size (Wardlaw *et al*., 2011; Deary *et al*., 2012; Taylor, Pattie and Deary, 2018; Stevenson *et al*., 2020; Conole *et al*., 2021).

Ethical approval for the LBC1936 study was obtained from the Multi-Centre Research Ethics Committee for Scotland (MREC/01/0/56) and the Lothian Research Ethics Committee (LREC/2003/2/29) for Wave 1, and the Scotland A Research Ethics Committee (07/MRE00/58) for Waves 2–5. All participants provided written informed consent.

### 2.2 Depression phenotypes

#### Generation Scotland

Lifetime MDD case/control status was ascertained using the research version of the Structured Clinical Interview for DSM disorders (SCID) (First *et al*., 2002) and diagnostic criteria were based on the ‘Diagnostic and Statistical Manual of Mental Disorders’ (DSM-IV-TR). A continuous measure of depressive symptom severity was also collected using the ‘Quick Inventory of Depressive Symptomatology’ (QIDS) (John Rush *et al*., 1986) which is a 16-item questionnaire. Results for analyses using the continuous MDD symptom measure are provided in the supplementary materials (Table S3).

#### LBC1936

Depression was measured at wave 2 using the depression subscale of the Hospital Anxiety and Depression Scale (HADS-D) (Zigmond and Snaith, 1983). Participants were classified as depressed when they had a score of 8 or above on the HADS-D subscale in accordance with previous literature (Olssøn, Mykletun and Dahl, 2005). Additional analyses including the HADS-D as a continuous measure is provided in the supplementary materials (Table S3).

### 2.3 Serum C-reactive protein (CRP) measurement

#### Generation Scotland

Blood samples were collected by venepuncture, extracted in to clot activator gel for serum separation and sent to NHS laboratories (Aberdeen Royal Infirmary or Ninewells Hospital). A low-sensitivity assay was used to detect serum CRP levels with a detection threshold limit of 4mg/L. Based on this threshold limit, CRP levels were stratified into clinically relevant groups: <4mg/L (coded as 0) and ≥4mg/L (coded as 1).

#### LBC1936

Serum CRP was measured from whole-blood samples using a high sensitivity assay (enzyme-linked 308 immunosorbent assay; R&D Systems, Oxford, UK) (Aribisala *et al*., 2014). For analyses purposes the CRP levels were stratified in to <4mg/L (coded as 0) and ≥4mg/L (coded as 1) to be consistent with the GS-imaging sample. Additional analyses including serum CRP as a continuous variable is included in the supplementary materials (Table S4).

### 2.4 DNAm CRP score calculation

Full details of the DNAm CRP score calculation, methylation data pre-processing and quality control for both the GS-imaging and LBC1936 cohorts have been reported in full previously (Stevenson *et al*., 2020). For the GS-imaging sample, methylation data was collected at baseline and was therefore not concurrent with imaging assessment and so the time between methylation collection and brain imaging collection was included in analyses (Green, Shen, *et al*., 2021). For both the GS-imaging and LBC1936 cohorts, methylation beta values were extracted for 7 CpG sites with the strongest evidence of a functional association with serum CRP, as derived from a large sample with both European and African ancestries (Ligthart *et al*., 2016). One of the 7 CpG sites (cg06126421) was not available in the Generation Scotland cohort and therefore 6 CpG sites were included in the Generation Scotland analysis. All 7 CpG sites were available in the LBC1936 cohort. The DNAm CRP score with 6 CpGs was highly correlated with the score including 7 CpGs when calculated in the LBC1936 sample (r=0.94, p<0.001). To calculate the DNAm CRP score, the beta values for the CpG sites were multiplied by their regression weights reported in the Ligthart et al study (Ligthart *et al*., 2016) and then summed to generate a DNAm CRP score for each participant.

### 2.5 MRI acquisition

#### Generation Scotland

Scanning took place at two study sites: Aberdeen and Dundee. Both study sites followed the same protocol, structural sequences and acquisition parameters, which have been described in full previously (Habota *et al*., 2019; Green, Shen, *et al*., 2021). Briefly, Aberdeen participants were scanned using a 3T Philips Achieva TX-series MRI system (Philips Healthcare, Best, Netherlands) with a 32-channel phased-array head coil and a back-facing mirror (software version 5.1.7; gradients with maximum amplitude 80 mT/m and maximum slew rate 100 T/m/s). In Dundee participants were scanned with a Siemens 3T Prisma-FIT (Siemens, Erlangen, Germany) with 20-channel head and neck phased array coil and a back-facing mirror (Syngo E11, gradient with max amplitude 80 mT/m and maximum slew rate 200 T/m/s) (Habota *et al*., 2019).

#### LBC1936

Full MRI acquisition parameters for the LBC1936 cohort have been described previously (Wardlaw *et al*., 2011). Briefly, structural MRI acquisition and processing in LBC1936 were performed according to an open-access protocol with a 1.5 T GE Signa HDx clinical scanner (General Electric, Milwaukee, WI, USA) which was used to collect structural T1-, T2-, T2*-weighted, and fluid attenuated inversion recovery (FLAIR) images.

### 2.6 Brain age calculation

For both cohorts, brain age was calculated using ‘brainageR’ (version 2.1 available at https://github.com/james-cole/brainageR). BrainageR predicts brain age using a machine learning algorithm and utilises voxel-wise normalised grey matter, white matter and CSF volumetric data derived from T1-weighted MRI (Cole *et al*., 2018). The calculated brain age variable was then residualised over age and sex for both cohorts and additionally scan site for the GS-imaging sample to produce a measure of brain-predicted age difference (brain-PAD).

### 2.7 White matter hyperintensity phenotypes

For both cohorts, WMH scores were derived for each participant using the Fazekas and Wahlund visual scales (Fazekas *et al*., 1987; Wahlund *et al*., 2001) by a trained observer (EB, ZM, checked by JMW) blind to clinical and all other information. Using the Fazekas scale, WMH were defined as punctuate, focal or diffuse lesions in the deep or periventricular white matter, basal ganglia or brainstem, where they appeared as visible areas of hyperintensity on the FLAIR images (Shi *et al*., 2021). Severity of WMH was graded as 0 (absent) to 3 (severe) for periventricular and deep WMH. A score of 1 is defined as caps or pencil thin lining or punctuate foci for periventricular and deep WMH respectively. A score of 2 is defined as a smooth halo or beginning to confluence. A score 3 is defined as an irregular periventricular signal extending into the deep white matter or large confluent areas. A total Fazekas score (0-6) was calculated for analyses purposes by summing periventricular and deep WMH scores (0-3 each). Overall, this provided three phenotypes for both cohorts (Fazekas deep white matter, Fazekas periventricular and Fazekas total).

The Wahlund scale ranges from 0 to 30 and scores WMH on the scale 0 to 3 in five regions of the brain on both left and right sides: frontal, parieto-occipital, temporal, infratentorial/cerebellum and basal ganglia (Wahlund *et al*., 2001). A score of 0 represents no lesions and a score of 3 represents “diffuse involvement of the entire region” and in the basal ganglia represents confluent lesions. This provided five white matter hyperintensity phenotypes for both cohorts for the five brain areas scored in the Wahlund scale.

### 2.8 Statistical analyses

All statistical analyses were conducted using R (version 4.1.0) and the maximum available size was used in all analyses. Generalised linear models (function ‘glm’ in R package ‘stats’) were used to investigate the association between inflammatory markers and depression with markers of brain ageing. False Discovery Rate (FDR) correction for multiple testing was applied, referred to as P_FDR_ in this report, using the ‘p.adjust’ function in R (Benjamini and Hochberg, 1995). All betas were standardised. Covariates for MDD analyses in both cohorts included age, sex and additionally study site for the GS-imaging cohort. Covariates for serum CRP analyses in both cohorts included age, sex, body mass index, smoking status and additionally study site for the GS-imaging cohort. Covariates for DNAm CRP analyses in both cohorts were the same as serum CRP but additionally included methylation set and for the GS-imaging cohort, the time between methylation data collection and the neuroimaging appointment. Recent work has examined associations between MDD and brain-PAD in these data (Harris *et al*., 2021: in preparation). Here, we investigated: (i) associations between serum CRP and DNAm CRP with brain-PAD, (ii) associations between serum CRP and DNAm CRP with WMH phenotypes, (iii) associations between MDD status and WMH phenotypes and (iv) interaction effects between these inflammatory makers and MDD on brain-PAD/WMH measures.

## 3. Results

### 3.1 Cohort demographics

Descriptive statistics of the key variables for each cohort are presented in Table 1. Briefly, participants in the Generation Scotland sample (*N*=804) were younger (M= 59.5, SD=9.7) than the Lothian Birth Cohort participants (*N*=615, M=72.6, SD=0.7). The Generation Scotland sample also had more females (58.2%) compared to the Lothian Birth Cohort (46.7%).

### 3.2 Inflammatory markers and brain age

We first tested the association between the two markers of inflammation (serum CRP and DNAm CRP) and brain-PAD. Serum CRP was not significantly associated with brain-PAD in either cohort (β_GS_= -0.027, p_GS_=0.503; β_LBC1936_=0.032, p_LBC1936_=0.451, Figure 1). However, in both cohorts a greater DNAm CRP score was associated with a higher (older-appearing) brain-PAD. As shown in Figure 2, the magnitude of the effect was similar in both the GS-imaging sample (β=0.111, p=0.015; equivalent to 0.7 years per standard deviation increase of DNAm CRP scores) and in the LBC1936 sample (β= 0.114, p=0.012; here equivalent to 0.75 years per standard deviation increase in DNAm CRP). There were no differences in the pattern of results between using serum CRP stratified (<4mg/L and ≥4mg/L) or as a continuous variable for the LBC1936 analyses (Table S1 and S4).

**Figure 1:**
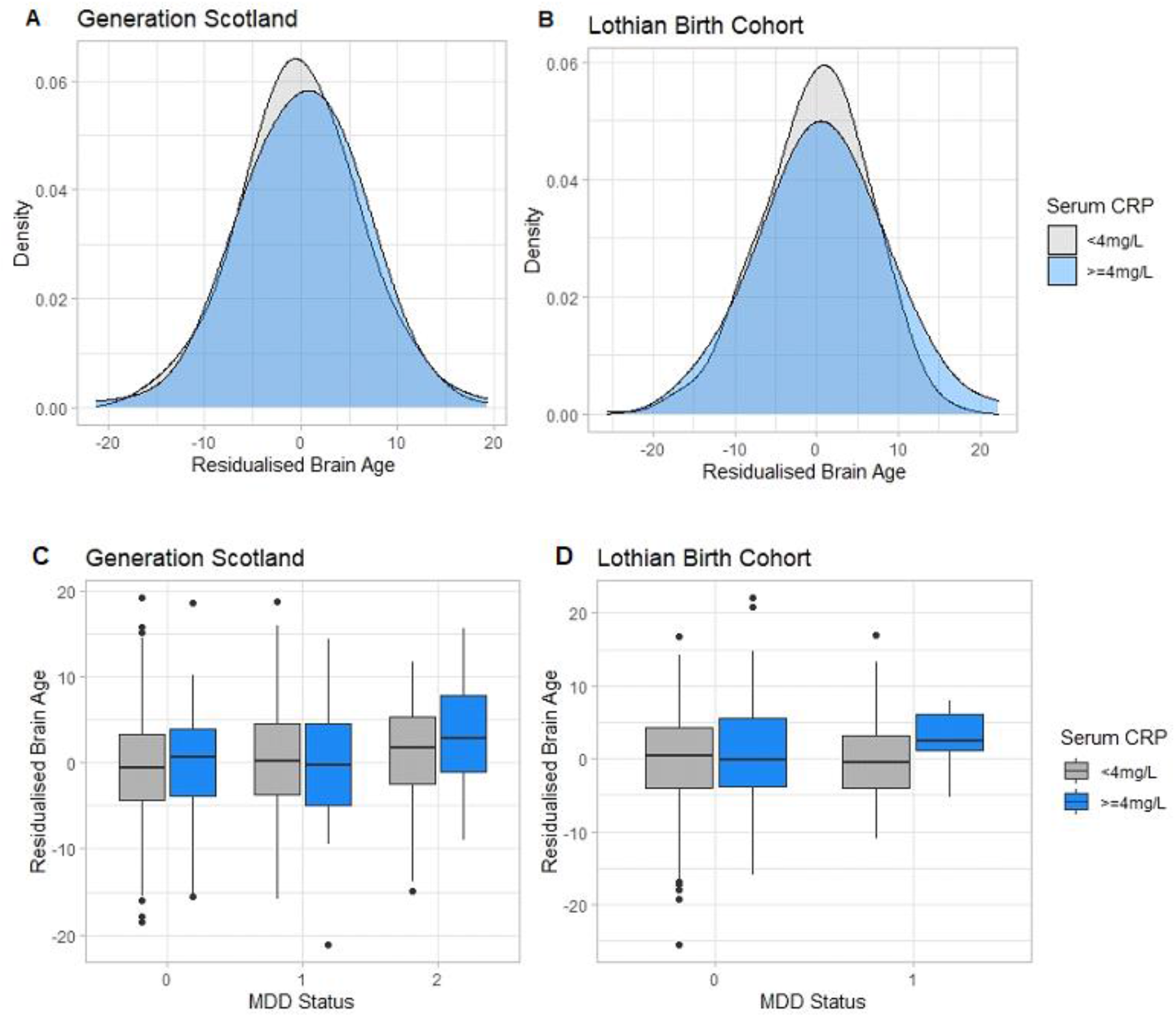
The association between (**A**) serum CRP status and brain age (brain-PAD) in Generation Scotland, (**B**) serum CRP status and brain-PAD in LBC1936, (**C**) serum CRP and brain-PAD stratified by depression status in Generation Scotland and (**D**) serum CRP and brain-PAD stratified by depression status in LBC1936. (**A and B**) Serum CRP status and residualised brain age (brain-PAD). (**C**) MDD status is stratified as 0=no history of MDD, 1=previous episode of MDD and 2=current episode of MDD using SCID diagnoses. (**D**) MDD status is stratified as 0= no MDD and 1=current MDD using HADS-D scores. In all plots, serum CRP status is stratified as 0=<4mg/L and 1 >=4mg/L and brain age (brain-PAD) is residualised over age, sex and scan site and scaled and centred on the mean.

**Figure 2:**
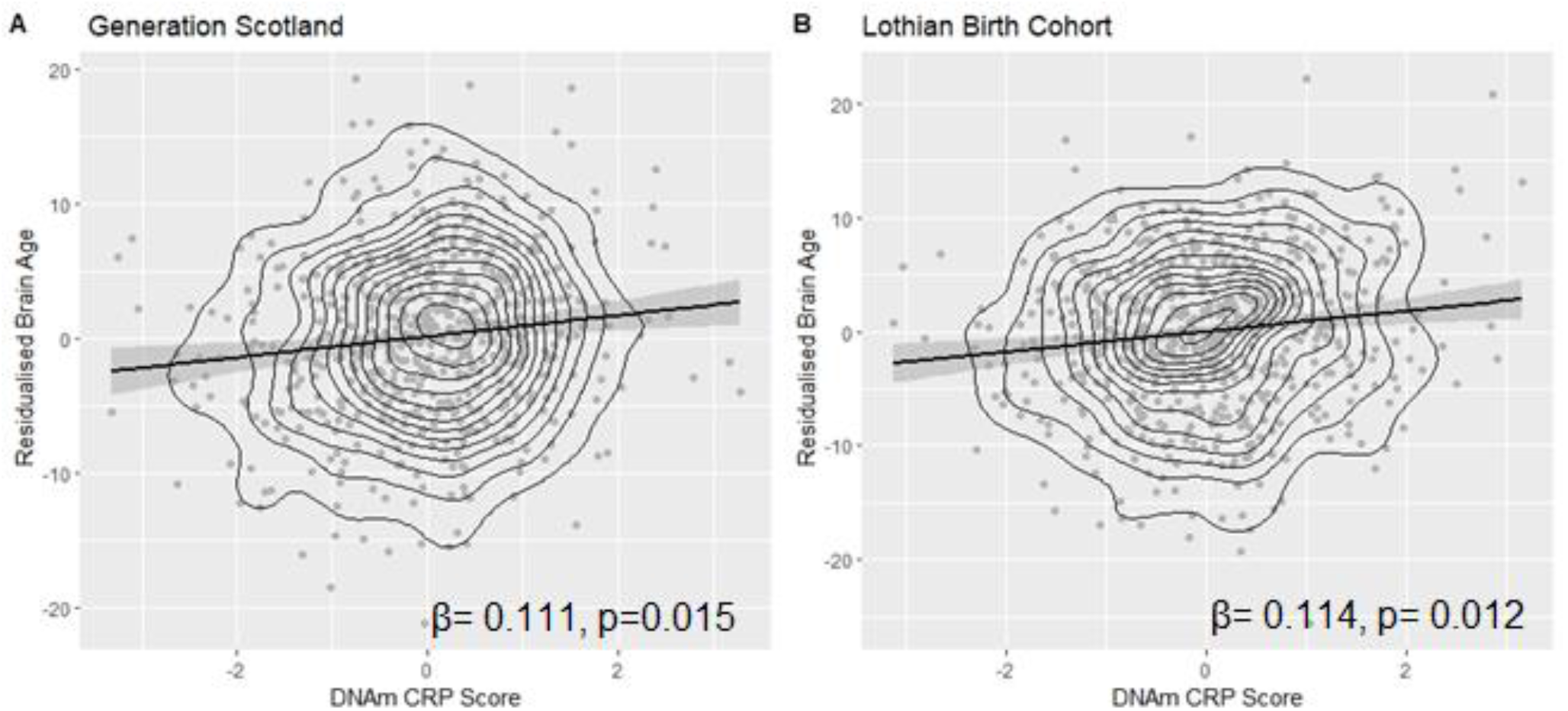
Association between DNAm CRP and brain age (brain-PAD) in the (**A**) Generation Scotland sample and (**B**) LBC1936 cohort. Brain age (brain-PAD) is residualised over age, sex and scan site. Both brain age and DNAm CRP are scaled and centred on the mean.

### 3.3 Inflammatory markers and white matter hyperintensities

Serum CRP status and DNAm CRP scores were not significantly associated with any of the WMH measures in the GS-imaging cohort, however, the effect sizes largely went in a positive direction (Table S1, Figure 3). In the LBC1936 cohort, while serum CRP was not associated with any WMH measure, DNAm CRP was associated with increased deep WMH (as measured by the Fazekas scale; β=0.116, P_FDR_=0.033) and increased total Fazekas scores (β=0.107, P_FDR_=0.033). The DNAm CRP score was also significantly associated with increased Wahlund infratentorial WMH scores in the LBC1936 cohort (β=0.15, P_FDR_=0.006).

**Figure 3:**
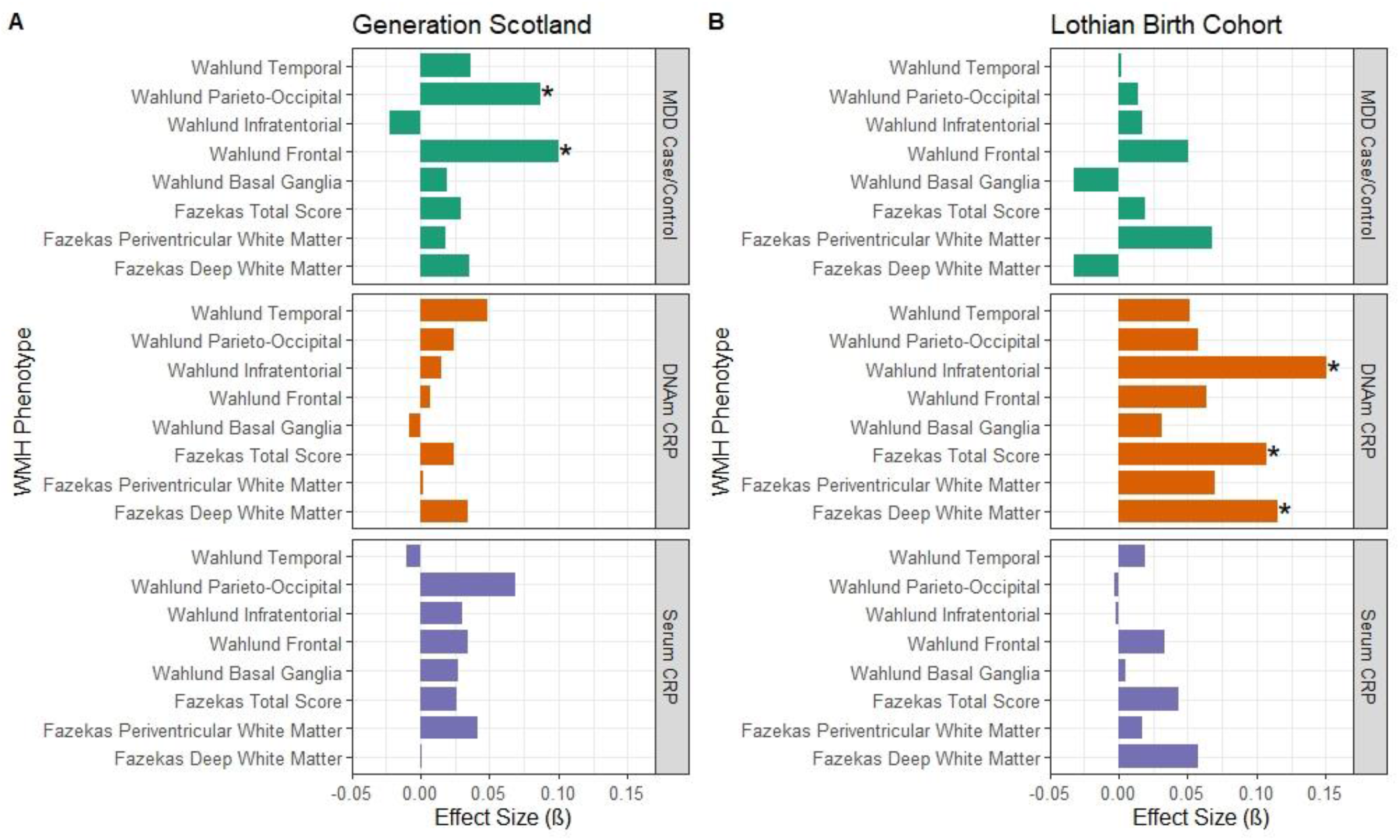
Standardised effect sizes for the association between MDD and inflammatory measures with white matter hyperintensity phenotypes in (**A**) Generation Scotland and (**B**) Lothian Birth Cohort. Associations where p=<0.05 are marked with an asterix. MDD case/control status is assessed by SCID diagnosis in the GS-imaging sample and by the HADS-D scale in the LBC1936 cohort.

### 3.4 Depression and white matter hyperintensities

MDD case/control status was significantly associated with increased Wahlund scores in the frontal (β=0.1, P_FDR_=0.032) and parieto-occipital areas (β=0.087, P_FDR_=0.045) in the GS-imaging cohort. Details of further analyses in the GS-imaging cohort looking at other features of MDD (recurrence, age of onset and number of episodes) and associations with WMH measures are provided in the supplementary materials, although no associations survived FDR correction (Table S5). In the LBC1936 cohort, there was no significant relationship between WMH and MDD (as measured by HADS-D scores). There were also no differences in the pattern of results using HADS-D stratified (<8= no MDD, ≥8 = MDD) or as a continuous variable (Table S3).

### 3.5 Interaction effects between inflammation markers and depression on brain ageing phenotypes

We tested the interaction effects between both inflammatory markers (serum CRP and DNAm CRP) and MDD on brain-PAD. There was no significant interaction effect between serum CRP and MDD status (β= -0.052, p=0.505), nor between DNAm CRP and MDD case/control status on brain-PAD in the GS-imaging cohort (β=0.064, p=0.654). There were also no significant interaction effects between either inflammatory marker (serum CRP and DNAm CRP) and the measure of MDD (HADS-D) on brain-PAD in the LBC1936 cohort.

There were no significant interaction effects between either marker of inflammation (serum CRP/ DNAm CRP) with MDD measures on any of the WMH measures (all P_FDR_>0.05) in either cohort (Table S2).

## 4. Discussion

To our knowledge this is the first study to investigate a DNAm marker of inflammation and its associations with markers of brain ageing in the context of depression. This study utilised a large and well-phenotyped cohort of individuals in the Generation Scotland dataset and we sought replication in another large independent sample - the Lothian Birth Cohort 1936. We show that a methylation-derived score of CRP was significantly associated with an increased brain-PAD (an older-looking brain relative to chronological age), and that the direction and magnitude of the effect replicated in an independent sample. DNAm CRP was further associated with more pronounced WMHs in the older LBC1936 sample. Serum CRP was not associated with any marker of brain ageing in this study in either cohort, demonstrating the utility of methylomic inflammatory profiles which are more stable longitudinally and may capture longer-term inflammatory associations (Stevenson *et al*., 2020; Conole *et al*., 2021; Green, Shen, *et al*., 2021). MDD case/control status was associated with WMH markers of brain ageing in the frontal and parieto-occipital areas in the GS-imaging cohort, however this was not replicated in the LBC1936 sample. There were also no interaction effects between inflammation and MDD on brain ageing phenotypes in either cohort. Taken together, these findings demonstrate that both inflammation and MDD may be separately associated with differing markers of brain ageing (brain-PAD and WMH respectively). However, there was an absence of an interaction between MDD status and inflammatory markers on brain ageing, indicating that the ‘inflammaging’ effects seen were not specific to MDD in these relatively healthy community-dwelling samples.

The most robust finding in this study however was the significant association between DNAm CRP scores and increased brain ageing that was replicated in an independent sample. Brain ageing acceleration was equivalent to an increase in 0.7 years per standard deviation of the DNAm CRP score in the GS-imaging sample and 0.75 years in the LBC1936 cohort. These findings build upon previous work that has reported that DNAm CRP scores are associated with widespread deficits in white matter structural connectivity and poor brain health outcomes, suggesting that chronic, longer-term inflammation does indeed have ‘inflammaging’ effects on the brain (Conole *et al*., 2021; Green, Shen, *et al*., 2021). The results are also consistent with previous research that has shown that chronic inflammation is associated with neurodegenerative phenotypes and cognitive decline (Engelhart *et al*., 2004; Tilvis *et al*., 2004; Matthews, 2019; Stevenson *et al*., 2020; Conole *et al*., 2021). In the older LBC1936 cohort, the DNAm CRP score was further associated with increased WMH in the deep white matter and infratentorial regions. This is line with previous research that has found that systemic inflammation is associated with WMH deterioration and proliferation and that prolonged inflammation is a key contributor to white matter pathology (Raz *et al*., 2012; Walker *et al*., 2017). Overall, the current study suggests there may be a chronic inflammatory component to advanced brain ageing in terms of both calculated brain age and white matter disease. However, the direction of causality is unknown at present and methods such as Mendelian randomisation would be required to attempt to address the causal nature of the observed associations in this study.

We further report that MDD status was also associated with brain ageing phenotypes, specifically increased WMH in the frontal and parieto-occipital regions. This is consistent with a large body of previous evidence demonstrating that MDD, in particular late-life depression, is associated with increased WMHs, thought to be ischemic/ cerebrovascular in origin (Firbank et al. 2005; Herrmann, Le Masurier, and Ebmeier 2008; Taylor et al. 2005). The frontal lobe is also important for cognition and emotional regulation and pathology in this area may explain the emotional dysregulation and cognitive dysfunction seen in MDD (de Nooij *et al*., 2020). These findings were however not replicated in the LBC1936 sample. However, clinically ascertained MDD case/control status was not available in the older LBC1936 cohort (where a HADS-D cut off was used). Further, only a small number of individuals in the LBC1936 cohort met criteria for MDD (*N*=26). It is therefore possible that differences between the cohorts may contribute to lack of replication here, including the differences in MDD classification. The LBC1936 sample was also much older than the GS-imaging sample, with an increased likelihood of age-related WMHs, which may potentially obscure depression-related WMH associations present at younger ages.

Inflammation has been previously linked to somatic symptoms of MDD (Iob, Kirschbaum and Steptoe, 2020; Green, Shen, *et al*., 2021) and previous research has found that advanced brain ageing in MDD was most strongly associated with somatic depressive symptomatology (Han *et al*., 2021). However, we found no evidence of interaction effects between inflammation and MDD with any of the brain ageing measures in this study. There are several potential reasons for this lack of association that should be considered in the interpretation of these findings. Firstly, both samples were community-based which may have introduced sampling bias as there were few currently unwell individuals and few with severe MDD. This means that this study is potentially underpowered to detect associations with current and moderate-severe MDD, for example in individuals who require hospitalisation. Furthermore, work has demonstrated that the range restriction in the LBC1936 sample means it is likely that effect sizes are underestimates of those in the general population (Johnson *et al*., 2011).

There are further limitations of this study to consider. The serum CRP measure in the GS-imaging sample was conducted using a low sensitivity assay, with a detection threshold of 4mg/L. However, we note that in the LBC1936 sample, that did have a high sensitivity measure, the results were replicated lending confidence to these findings. The DNAm CRP measure in the GS-imaging cohort was also collected at baseline appointment and was not concurrent with imaging, however, we did account for this time difference in the analyses. The LBC1936 DNAm CRP measure was also concurrent with neuroimaging and replicated the GS-imaging findings, demonstrating the validity of these results. There are also important demographic differences to consider between the two cohorts in this study. The LBC1936 sample additionally had a restricted age range of individuals as all of the LBC1936 participants were approximately 73 years of age. However, this removes the strong confounding effects of age and cohort effects of wider age range studies.

This study also has several strengths, in particular, the use of two large independent cohorts (*N*_GS_=804, *N*_LBC1936_=615) with in-depth phenotypic data integrating clinical, epigenetic and neuroimaging data. Our methylation-based measure of longer-term inflammatory activity is more temporally stable compared to cross-sectional serum measures and may represent a longitudinal archive of previous inflammatory exposure (Stevenson *et al*., 2020). Our study thus provides a powerful example of the utility of methylomic profiles in the investigation of MDD and brain phenotypes. Future research should investigate MDD, inflammation and brain ageing processes in larger cohorts with a greater number of individuals with current and clinically ascertained symptoms to disentangle these relationships further. In addition to DNAm CRP, future studies could explore methylation signatures of further inflammatory mediators including proinflammatory cytokines such as interleukin-6 to provide further insight in to the role of inflammatory mechanisms in MDD and brain ageing (Stevenson *et al*., 2021).

In conclusion, this study utilised two large independent cohorts to investigate the role of inflammation in brain ageing and MDD phenotypes. We found a robust association between a temporally stable and cumulative epigenetic inflammation score (DNAm CRP) and advanced brain ageing that was replicated in an independent cohort. This longer-term epigenetic marker of inflammation was also associated with white matter pathology in the older cohort, indicating ‘inflammaging’ effects on the brain. We also report that MDD was associated with accelerated brain ageing as measured by white matter pathology in the MDD focused cohort, with increased WMH in the frontal lobe, which is important for cognition and emotion regulation. However, we did not find any evidence that observed associations between brain ageing markers and inflammation were stronger in those with MDD relative to controls, indicating no evidence of an elevated liability of inflammaging in depression. Taken together, these findings indicate that inflammation and MDD are both independently associated with brain ageing phenotypes, but the current results did not indicate that inflammation was a key component of the brain ageing seen in MDD in our samples. Future research should aim to investigate these relationships in clinical rather than community-based samples, explore further inflammatory markers and characterise the potential effects of inflammatory mechanisms on MDD brain pathology and symptomatology.

## Data Availability

Access to and use of GS data must be approved by the GS Access Committee under the terms of consent. Full details of the application process can be found at www.generationscotland.org. LBC1936 data can be requested via a data access request to the Lothian Birth Cohorts research group at www.ed.ac.uk/lothian-birth-cohorts/data-access-collaboration.

## Acknowledgments

This research was funded in whole or in part by the Wellcome Trust (grant numbers 104036/Z/14/Z; 221890/Z/20/Z; 216767/Z/19/Z). For the purpose of open access, the author has applied a CC-BY public copyright license to any author-accepted manuscript version arising from this submission.

The authors would like to thank all of the Generation Scotland and Lothian Birth Cohort 1936 participants for their contribution to this study. Generation Scotland received core support from the Chief Scientist Office of the Scottish Government Health Directorates [CZD/16/6] and the Scottish Funding Council [HR03006] and is currently supported by the Wellcome Trust [216767/Z/19/Z]. Genotyping of the GS:SFHS samples was carried out by the Genetics Core Laboratory at the Edinburgh Clinical Research Facility, University of Edinburgh, Scotland and was funded by the Medical Research Council UK and the Wellcome Trust (Wellcome Trust Strategic Award “STratifying Resilience and Depression Longitudinally” (STRADL) Reference 104036/Z/14/Z).

The LBC1936 research is supported by Age UK (Disconnected Mind project) and by the UK Medical Research Council [MRC; G0701120, G1001245, MR/M013111/1, MR/K026992/1], and the University of Edinburgh. The Lothian Birth Cohort 1936 study acknowledges the financial support of NHS Research Scotland (NRS) through the Edinburgh Clinical Research Facility.

SRC is supported by a Sir Henry Dale Fellowship, jointly funded by the Wellcome Trust and the Royal Society (Grant Number 221890/Z/20/Z). JHC is supported by a UKRI Innovation Fellowship (MR/R024790/2). CG is supported by The Medical Research Council and The University of Edinburgh through the Precision Medicine Doctoral Training program.

