## Supplementary_Material for "‘Inflammaging’ in Major Depressive Disorder: Associations between markers of inflammation and brain ageing in depression across two large community-based cohorts"

**[Supplementary Materials]**

**WMH Volume and Count Measures in Generation Scotland**

In the Generation Scotland sample, two additional WMH-related phenotypes were derived using the Lesion Segmentation Toolbox (LST) version 3.0 for SPM (<https://www.applied-statistics.de/lst.html>). The two measures were total WMH volume and WMH count and were obtained with the lesion growth algorithm (LGA). Results of analyses of these measures are included in Tables S1, S2, S3 and S5.

The LGA first segments the T1 images into the three main tissue classes (cerebrospinal fluid, grey matter and white matter) and this information is combined with FLAIR intensities to calculate lesion belief maps. These maps are thresholded with a pre-chosen initial threshold (κ parameter) to obtain an initial binary lesion map. This map is then grown along voxels that appear hyperintense in the FLAIR image, which produces a lesion probability map. The lesion probability map is then again thresholded to define WMH regions. The initial threshold (κ parameter) was first set to 0.3 for all participants, following past studies with the LGA (1,2). Lesion probability maps were then reviewed manually and for each participant all horizontal slices containing either missed WMH regions or WMH regions detected in error were recorded. For participants who had more than 5 slices with missed WMH, κ parameter was decreased to 0.2 and lesion segmentation was rerun. Accordingly, for participants who had more than 5 slices with WMH regions detected in error, κ parameter was increased to 0.4 and lesion segmentation was rerun. The rerun threshold of five slices was selected because this represents approximately at least 5% of all slices with white matter prone to hyperintensities. For participants where substantial WMH regions were still missed after the rerun, κ parameter was further decreased to 0.1 and lesion segmentation was rerun the second time. Final lesion probability maps were thresholded with the default value of 0.5 to calculate WMH counts and total WMH volumes for each participant.

**Table S1:** Inflammation associations with brain ageing phenotypes in the Generation Scotland and Lothian Birth Cohort samples.

| **Brain Ageing Phenotype** | **Generation Scotland- Imaging** | | | **Lothian Birth Cohort** | | |
| --- | --- | --- | --- | --- | --- | --- |
|  | **β** | **SE** | **pFDR** | **β** | **SE** | **pFDR** |
| **Serum CRP Results** |  |  |  |  |  |  |
| Brain-PAD | -0.027 | 0.039 | 0.503 | 0.032 | 0.042 | 0.451 |
| Fazekas Periventricular White Matter | 0.041 | 0.038 | 0.707 | 0.017 | 0.042 | 0.682 |
| Fazekas Deep White Matter | 0.001 | 0.037 | 0.987 | 0.058 | 0.041 | 0.449 |
| Fazekas Total | 0.026 | 0.037 | 0.707 | 0.043 | 0.042 | 0.449 |
| Wahlund Frontal | 0.034 | 0.040 | 0.606 | 0.033 | 0.042 | 0.952 |
| Wahlund Parieto-Occipital | 0.069 | 0.040 | 0.426 | -0.003 | 0.042 | 0.952 |
| Wahlund Temporal | -0.010 | 0.040 | 0.803 | 0.019 | 0.041 | 0.952 |
| Wahlund Infratentorial | 0.030 | 0.040 | 0.606 | -0.003 | 0.042 | 0.952 |
| Wahlund Basal Ganglia | 0.027 | 0.039 | 0.606 | 0.005 | 0.042 | 0.952 |
| WMH Volume Total | -0.012 | 0.032 | 0.969 | - | - | - |
| WMH Count | -0.001 | 0.035 | 0.969 | - | - | - |
| **DNAm CRP Results** |  |  |  |  |  |  |
| Brain-PAD | 0.111 | 0.046 | **0.015** | 0.114 | 0.046 | **0.012** |
| Fazekas Periventricular White Matter | 0.002 | 0.045 | 0.965 | 0.070 | 0.047 | 0.135 |
| Fazekas Deep White Matter | 0.034 | 0.044 | 0.867 | 0.116 | 0.046 | **0.033** |
| Fazekas Total | 0.024 | 0.043 | 0.867 | 0.107 | 0.047 | **0.033** |
| Wahlund Frontal | 0.007 | 0.047 | 0.887 | 0.064 | 0.047 | 0.340 |
| Wahlund Parieto-Occipital | 0.024 | 0.047 | 0.887 | 0.058 | 0.047 | 0.340 |
| Wahlund Temporal | 0.049 | 0.047 | 0.887 | 0.051 | 0.047 | 0.340 |
| Wahlund Infratentorial | 0.015 | 0.047 | 0.887 | 0.150 | 0.046 | **0.006** |
| Wahlund Basal Ganglia | -0.008 | 0.044 | 0.887 | 0.031 | 0.047 | 0.507 |
| WMH Volume Total | 0.055 | 0.038 | 0.269 | - | - | - |
| WMH Count | 0.044 | 0.040 | 0.269 | - | - | - |

**Table S2:** Inflammation x MDD interaction effects on brain ageing phenotypes in the Generation Scotland and Lothian Birth Cohort samples.

| **Brain Ageing Phenotype x MDD** | **Generation Scotland- Imaging** | | | **Lothian Birth Cohort** | | |
| --- | --- | --- | --- | --- | --- | --- |
|  | **β** | **SE** | **pFDR** | **β** | **SE** | **pFDR** |
| **Serum CRP x MDD Results** |  |  |  |  |  |  |
| Brain-PAD | -0.052 | 0.077 | 0.505 | 0.091 | 0.170 | 0.621 |
| Fazekas Periventricular White Matter | 0.103 | 0.076 | 0.249 | 0.247 | 0.170 | 0.444 |
| Fazekas Deep White Matter | 0.083 | 0.072 | 0.249 | -0.047 | 0.169 | 0.782 |
| Fazekas Total | 0.112 | 0.072 | 0.249 | 0.112 | 0.170 | 0.766 |
| Wahlund Frontal | 0.118 | 0.079 | 0.678 | 0.253 | 0.169 | 0.343 |
| Wahlund Parieto-Occipital | 0.065 | 0.078 | 0.685 | 0.137 | 0.170 | 0.526 |
| Wahlund Temporal | 0.067 | 0.078 | 0.685 | -0.250 | 0.168 | 0.343 |
| Wahlund Infratentorial | 0.002 | 0.078 | 0.982 | 0.101 | 0.170 | 0.555 |
| Wahlund Basal Ganglia | 0.039 | 0.077 | 0.770 | 0.149 | 0.172 | 0.526 |
| WMH Volume Total | 0.110 | 0.063 | 0.167 | - | - | - |
| WMH Count | 0.083 | 0.069 | 0.227 | - | - | - |
| **DNAm CRP x MDD Results** |  |  |  |  |  |  |
| Brain-PAD | 0.064 | 0.093 | 0.654 | 0.021 | 0.255 | 0.935 |
| Fazekas Periventricular White Matter | 0.041 | 0.090 | 0.650 | -0.025 | 0.257 | 0.921 |
| Fazekas Deep White Matter | 0.110 | 0.087 | 0.531 | -0.058 | 0.255 | 0.921 |
| Fazekas Total | 0.081 | 0.087 | 0.531 | -0.048 | 0.256 | 0.921 |
| Wahlund Frontal | 0.087 | 0.094 | 0.587 | 0.027 | 0.256 | 0.947 |
| Wahlund Parieto-Occipital | 0.038 | 0.094 | 0.697 | -0.165 | 0.258 | 0.881 |
| Wahlund Temporal | 0.036 | 0.093 | 0.697 | -0.163 | 0.258 | 0.881 |
| Wahlund Infratentorial | 0.129 | 0.093 | 0.587 | -0.017 | 0.254 | 0.947 |
| Wahlund Basal Ganglia | 0.086 | 0.089 | 0.587 | -0.597 | 0.257 | 0.102 |
| WMH Volume Total | -0.017 | 0.076 | 0.818 | - | - | - |
| WMH Count | 0.038 | 0.081 | 0.818 | - | - | - |

**Table S3:** MDD associations with white matter hyperintensity phenotypes in the Generation Scotland and Lothian Birth Cohort samples.

| **Brain Ageing Phenotype** | **Generation Scotland- Imaging** | | | **Lothian Birth Cohort** | | |
| --- | --- | --- | --- | --- | --- | --- |
|  | **β** | **SE** | **pFDR** | **β** | **SE** | **pFDR** |
| **MDD Stratified** |  |  |  |  |  |  |
| Fazekas Periventricular White Matter | 0.018 | 0.036 | 0.604 | 0.067 | 0.041 | 0.292 |
| Fazekas Deep White Matter | 0.035 | 0.034 | 0.597 | -0.033 | 0.040 | 0.628 |
| Fazekas Total | 0.029 | 0.034 | 0.597 | 0.019 | 0.040 | 0.639 |
| Wahlund Frontal | 0.100 | 0.037 | **0.032** | 0.051 | 0.040 | 0.900 |
| Wahlund Parieto-Occipital | 0.087 | 0.037 | **0.045** | 0.015 | 0.041 | 0.900 |
| Wahlund Temporal | 0.036 | 0.036 | 0.527 | 0.002 | 0.040 | 0.952 |
| Wahlund Infratentorial | -0.022 | 0.037 | 0.597 | 0.018 | 0.040 | 0.900 |
| Wahlund Basal Ganglia | 0.019 | 0.036 | 0.597 | -0.033 | 0.040 | 0.900 |
| WMH Volume Total | 0.054 | 0.036 | 0.258 | - | - | - |
| WMH Count | 0.009 | 0.032 | 0.774 | - | - | - |
| **MDD Continuous** |  |  |  |  |  |  |
| Fazekas Periventricular White Matter | 0.028 | 0.034 | 0.413 | 0.050 | 0.041 | 0.650 |
| Fazekas Deep White Matter | 0.027 | 0.033 | 0.413 | -0.004 | 0.040 | 0.913 |
| Fazekas Total | 0.036 | 0.033 | 0.413 | 0.026 | 0.040 | 0.788 |
| Wahlund Frontal | 0.096 | 0.035 | **0.036** | 0.052 | 0.040 | 0.498 |
| Wahlund Parieto-Occipital | 0.061 | 0.036 | 0.224 | 0.003 | 0.041 | 0.944 |
| Wahlund Temporal | 0.017 | 0.035 | 0.930 | 0.033 | 0.040 | 0.521 |
| Wahlund Infratentorial | -0.002 | 0.036 | 0.955 | 0.061 | 0.040 | 0.498 |
| Wahlund Basal Ganglia | 0.011 | 0.035 | 0.930 | 0.034 | 0.040 | 0.521 |
| WMH Volume Total | 0.043 | 0.035 | 0.221 | - | - | - |
| WMH Count | 0.060 | 0.031 | 0.107 | - | - | - |

*Notes:* In the GS sample, stratified MDD was defined according to SCID interview and the continuous measure of MDD was defined as QIDS scores. In LBC1936, stratified MDD was defined as HADS-D >= 8, and the continuous measure of MDD was defined by the HADS-D score itself.

**Table S4:** Associations of serum CRP and serum CRP x MDD interaction with brain-PAD and WMH measures in the Lothian Birth Cohort, with serum CRP as a continuous variable.

| **Brain Ageing Phenotype** | **Lothian Birth Cohort** | | |
| --- | --- | --- | --- |
|  | **β** | **SE** | **pFDR** |
| **Serum CRP Continuous** |  |  |  |
| Brain-PAD | 0.048 | 0.041 | 0.316 |
| Fazekas Periventricular White Matter | -0.015 | 0.041 | 0.729 |
| Fazekas Deep White Matter | -0.014 | 0.041 | 0.729 |
| Fazekas Total | -0.017 | 0.041 | 0.729 |
| Wahlund Frontal | -0.018 | 0.041 | 0.743 |
| Wahlund Parieto-Occipital | -0.032 | 0.041 | 0.743 |
| Wahlund Temporal | -0.025 | 0.041 | 0.743 |
| Wahlund Infratentorial | -0.032 | 0.041 | 0.743 |
| Wahlund Basal Ganglia | -0.014 | 0.042 | 0.743 |
| **Serum CRP x MDD Continuous** |  |  |  |
| Brain-PAD | -0.009 | 0.013 | 0.498 |
| Fazekas Periventricular White Matter | -0.004 | 0.013 | 0.780 |
| Fazekas Deep White Matter | 0.010 | 0.013 | 0.780 |
| Fazekas Total | 0.004 | 0.013 | 0.780 |
| Wahlund Frontal | 0.000 | 0.013 | 0.971 |
| Wahlund Parieto-Occipital | 0.003 | 0.013 | 0.971 |
| Wahlund Temporal | -0.002 | 0.013 | 0.971 |
| Wahlund Infratentorial | -0.011 | 0.013 | 0.915 |
| Wahlund Basal Ganglia | -0.014 | 0.013 | 0.915 |

**Table S5:** Additional clinical MDD phenotypic associations with brain ageing phenotypes in the Generation Scotland cohort.

| **Brain Ageing Phenotype** | **Generation Scotland- Imaging** | | |
| --- | --- | --- | --- |
|  | **β** | **SE** | **pFDR** |
| **Recurrence** |  |  |  |
| Fazekas Periventricular White Matter | 0.040 | 0.065 | 0.813 |
| Fazekas Deep White Matter | -0.045 | 0.066 | 0.813 |
| Fazekas Total | 0.008 | 0.065 | 0.900 |
| Wahlund Frontal | 0.008 | 0.079 | 0.986 |
| Wahlund Parieto-Occipital | 0.055 | 0.078 | 0.986 |
| Wahlund Temporal | 0.013 | 0.066 | 0.986 |
| Wahlund Infratentorial | 0.045 | 0.059 | 0.986 |
| Wahlund Basal Ganglia | 0.001 | 0.063 | 0.986 |
| WMH Volume Total | -0.023 | 0.080 | 0.774 |
| WMH Count | -0.033 | 0.055 | 0.774 |
| **Number of Episodes (continuous)** |  |  |  |
| Fazekas Periventricular White Matter | 0.022 | 0.065 | 0.730 |
| Fazekas Deep White Matter | 0.064 | 0.067 | 0.615 |
| Fazekas Total | 0.054 | 0.065 | 0.615 |
| Wahlund Frontal | 0.107 | 0.080 | 0.515 |
| Wahlund Parieto-Occipital | -0.011 | 0.078 | 0.885 |
| Wahlund Temporal | 0.022 | 0.067 | 0.885 |
| Wahlund Infratentorial | 0.075 | 0.059 | 0.515 |
| Wahlund Basal Ganglia | -0.013 | 0.063 | 0.885 |
| WMH Volume Total | -0.018 | 0.080 | 0.818 |
| WMH Count | 0.020 | 0.056 | 0.818 |
| **Age of Onset (25 or less)** |  |  |  |
| Fazekas Periventricular White Matter | 0.026 | 0.068 | 0.697 |
| Fazekas Deep White Matter | -0.064 | 0.069 | 0.697 |
| Fazekas Total | -0.028 | 0.068 | 0.697 |
| Wahlund Frontal | -0.074 | 0.083 | 0.875 |
| Wahlund Parieto-Occipital | -0.031 | 0.081 | 0.875 |
| Wahlund Temporal | -0.011 | 0.069 | 0.875 |
| Wahlund Infratentorial | 0.021 | 0.062 | 0.875 |
| Wahlund Basal Ganglia | -0.067 | 0.066 | 0.875 |
| WMH Volume Total | 0.006 | 0.083 | 0.944 |
| WMH Count | -0.035 | 0.058 | 0.944 |

*Notes:* Recurrence is defined as presence of more than one episode of MDD, number of episodes was considered to be continuous, and age of onset was defined as a binary variable indicating MDD onset before or after 25 years of age.
